# A comparative mixed method study of flipped class room and traditional lecture for teaching rational drug prescription among undergraduates in Pharmacology

**DOI:** 10.1101/2023.02.17.23286082

**Authors:** Jitha Sushama, Scaria Thomas Pulikkunnel

## Abstract

Rational drug prescription is a very important competency an Indian Medical Graduate (IMG) should acquire during his undergraduate training. Irrational prescriptions lead to drug resistance, adverse drug reactions (ADRs) and drug interactions. This was a prospective interventional mixed – method study where students were pseudo-randomised to either Traditional Lecture or Flipped Classroom based on the date of their academic session. For the study purpose, 250 phase 2 MBBS students in Pharmacology were divided into two groups. Allocation into 2 groups was done by lot method one week before the scheduled class so that the online study materials could be sent to the students belonging to the flipped classroom method in the form of case scenarios and videos. A total of 198 students participated in the study with 103 in the TL group and 95 in the FC group. Acquisition of higher order cognitive skills like application and analysis was assessed by measuring an academic score obtained from a series of exercises on rational prescription in a given clinical scenario. The academic score of the TL group (Mean - 5.99, SD-2.34) vs FC (Mean-5.59, SD-1.39) group thus obtained was not statistically significant p > 0.05, Effect size-0.08. The perception of students towards TL and FC assessed with the help of a questionnaire showed a statistically significant difference in favour of FC for the perceptions in the ability to improve academic score, generate peer interaction, facilitate team work and improve teacher-student interaction.

## Introduction

Rational drug prescription is a very important competency an Indian Medical Graduate (IMG) should acquire during his undergraduate training. It has great impact on the health of the patients as well as community as a whole. Irrational prescriptions lead to drug resistance, adverse drug reactions (ADRs) and drug interactions. Proper training in writing prescriptions and rational use of drugs helps in improving patient safety, lessen the litigations on doctors, reduce the incidence of ADRs and drug interactions as well as build trust in the doctors.

Flipped classroom (FC) originated in American Woodland Park High School in 2007. The flipped or inverted classroom is a blended learning approach that reverses the traditional teaching^1^. When learning is flipped, didactic lectures are pre-recorded and made available for students to watch prior to class and class time is used for active learning strategies to deepen their understanding and knowledge^2^. The technological movement has enabled the amplification and duplication of information at an extremely low-cost^3^.

The effective delivery of a traditional lecture (TL) class poses a major challenge for the faculty members, especially in the field of medical education due to the involvement of vast knowledge and complicated concepts^4^. The flipped classroom has emerged as one of the most effective ways to impart curricular delivery during a large group teaching session^5^. The core elements of this learning include preclass content, assessments, working on gaps and developing competency. The teacher’s role is like a guide. It makes the real implementation of transformation from teacher-centered to student-centered. It makes the students from passive recipients of knowledge to active searcher for knowledge.

Some researches show that the flipped classroom method has a positive impact as an active learning method on various attitudes and behaviors such as student success, motivation, dependency, critical thinking, capability, creativity and problem-solving capability whereas some others have inconclusive results related to academic scores^6^. Even though it is quite an effective approach of teaching–learning, the implementation of the same requires careful planning and support from the faculty members^7^. FC has also been found to be advantageous in improvement of higher order cognitive skills like application, analysis, synthesis and evaluation.^8^

Literature search shows that there are very few studies which gauge student perception on multiple parameters as well as measure acquisition of higher order cognitive skills with FC versus TL in Pharmacology. The study aims to find the change in immediate outcomes as in the improvement in prescription writing and perception by students in the new teaching-learning method.

## Materials and Methods

### Study population

This was a prospective interventional mixed – method study where students were pseudo-randomised to either TL or FC based on the date of their academic session. Study was initiated after obtaining approval from the Institutional Human Ethics Committee (HEC No.01/17/2022/MCT dated 26/02/2022). Phase II MBBS students undergoing Pharmacology training willing to give informed consent were included in the study.

The study was conducted in the department of Pharmacology of a Government Medical College in South India.

### Study design

Prescriptions of the common disease conditions in general population as well as special groups like children and pregnant ladies are usually taught by lecture method. For the study purpose, 250 phase 2 MBBS students in Pharmacology were divided into two groups. Allocation into 2 groups was done by lot method one week before the scheduled class so that the online study materials could be sent to the students belonging to the flipped classroom method in the form of case scenarios and videos. First group was taught rational drug prescription by traditional lecture method and the second group was dealt with flipped classroom method. During the flipped classroom session student discussion was promoted to find the answers to the given clinical conditions. At the end of the session a short test in the form of Objective structured practical examination (OSPE) was conducted to write the prescription for a given clinical condition and the marks scored were assessed in both groups. A feedback form in the form of Likert questionnaire was given to the students after each session to assess how the students perceived each type of teaching method with respect to peer interaction, organization of topic, level of teacher student interaction, enhancing understanding of the topic, creating interest in the topic, improving communication skills, promoting team work, and developing motivation to learn rational prescription. The questionnaire also enquired if a teaching-learning method was expected to enhance their academic score and also if they would recommend the method to their juniors. The confidentiality of the students was maintained throughout the study about their feedback. The marks scored in the study was not used in the final assessment in internal or University examinations. Both the sessions were conducted by the same facilitator on consequent days.

To avoid ethical issues, lecture class was delivered at a later date for all students who had flipped classroom session.

### Statistical Analysis

The data collected using OSPE and feedback form were entered in Microsoft excel sheet and analysis done using SPSS software (IBM SPSS Statistics Version 20). The academic scores as well as student perception scored on a Likert scale were analysed using Mann Whitney U test. The perception of students obtained from feedback form were also expressed as percentages.

## Results

A total of 198 students participated in the study with 103 in the TL group and 95 in the FC group. Acquisition of higher order cognitive skills like application and analysis was assessed by measuring an academic score obtained from a series of exercises on rational prescription in a given clinical scenario. The academic score of the TL group (Mean - 5.99, SD-2.34) vs FC (Mean-5.59, SD-1.39) group thus obtained was not statistically significant p > 0.05, Effect size-0.08 The perception of students towards TL and FC were assessed with the help of a questionnaire. There was a statistically significant difference in favour of FC for the following perceptions: ability to improve academic score, generate peer interaction, facilitate team work and improve teacher-student interaction [Table 1].

**Figure 1:**
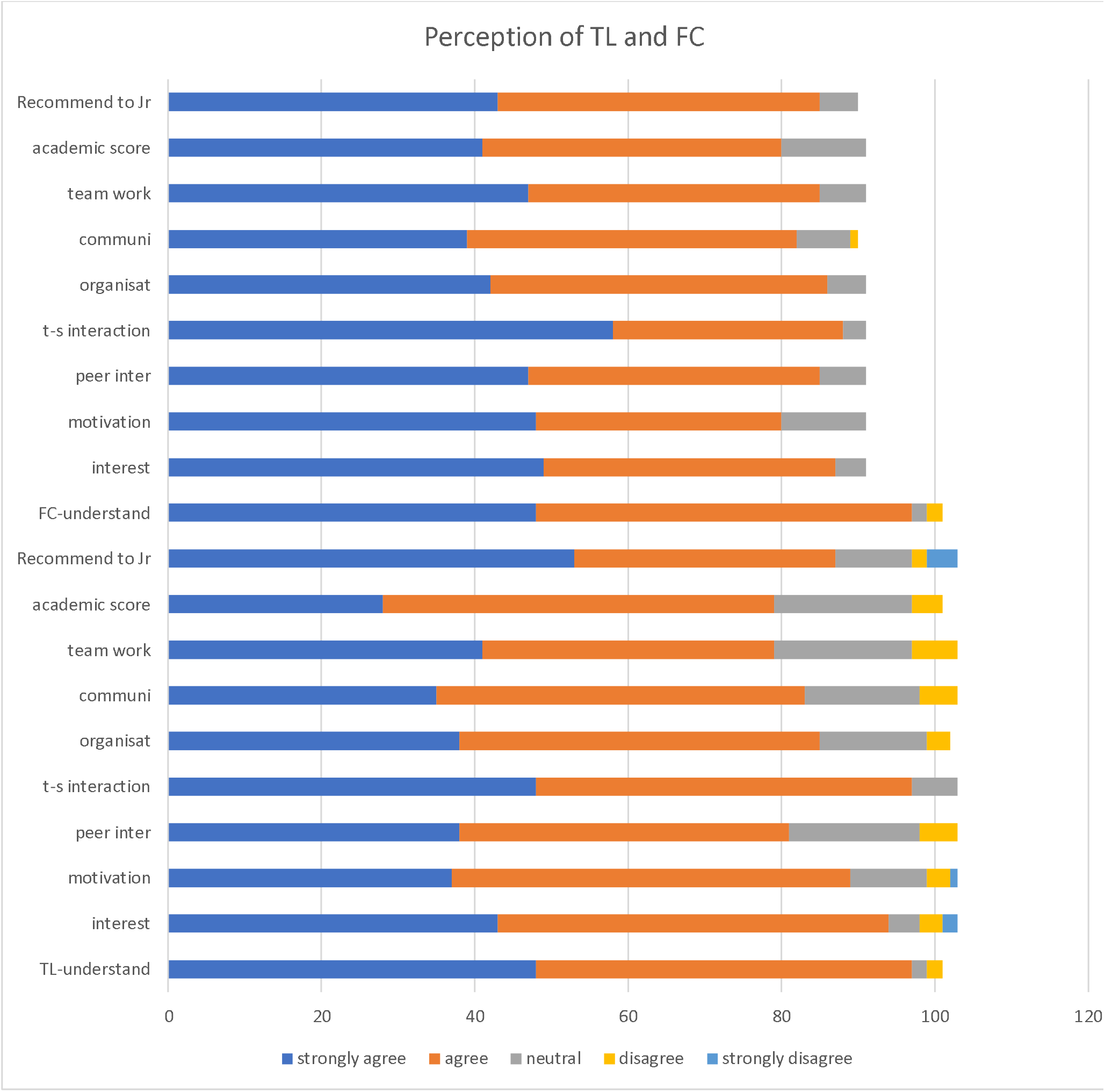
Perception of TL and FC.

**Table 1:** Mean perception score of TL compared to FC * In the Likert scale used in the study, 1= Strongly agree, 5= Strongly disagree. Hence lower the value more favourable the perception. #Effect size is calculated by test statistic divided by the root of sample size (small effect: 0.1 <r ≤ 0.3, medium effect: 0.3 <r ≤ 0.5, large effect: r > 0.5)

| Perception | Lecture |  | Flipped |  | P value | Effect size# |
| --- | --- | --- | --- | --- | --- | --- |
|  | Mean* | SD | Mean* | SD |  |  |
| Ability to improve academic score | 2.04 | 0.88 | 1.67 | 0.68 | 0.003 | 0.21 |
| Enhance communication skills | 1.90 | 0.82 | 1.66 | 0.67 | 0.054 | 0.14 |
| Create interest in topic | 1.74 | 0.83 | 1.50 | 0.58 | 0.064 | 0.13 |
| Increase motivation to learn | 1.83 | 0.79 | 1.66 | 0.69 | 0.176 | 0.09 |
| Better organisation of topic | 1.81 | 0.79 | 1.59 | 0.59 | 0.054 | 0.14 |
| Generate peer interaction | 1.89 | 0.85 | 1.55 | 0.62 | 0.005 | 0.2 |
| Promote team work | 1.89 | 0.89 | 1.55 | 0.62 | 0.01 | 0.18 |
| Recommend to juniors | 1.74 | 0.99 | 1.58 | 0.59 | 0.78 | 0.02 |
| Facilitate teacher student interaction | 1.59 | 0.60 | 1.39 | 0.55 | 0.016 | 0.17 |
| Greater understanding of topic | 1.55 | 0.67 | 1.49 | 0.52 | 0.45 | 0.05 |

**Table 2:** Distribution of Likert scale scores of perceptions

| Perception | Method | Likert Scale- No of responses (%) |  |  |  |  | Total responses |
| --- | --- | --- | --- | --- | --- | --- | --- |
|  |  | 1 | 2 | 3 | 4 | 5 |  |
| Ability to improve academic score | TL | 28<br>(27.2) | 51<br>(49.5) | 18<br>(17.5) | 4<br>(3.9) | 2<br>(1.9) | 103 |
|  | FC | 41<br>(45.1) | 39<br>(42.9) | 11<br>(12.1) | 0 | 0 | 91 |
| Enhance communication skills | TL | 35<br>(34) | 48<br>(46.6) | 15<br>(14.6) | 5<br>(4.9) | 0 | 103 |
|  | FC | 39<br>(43.3) | 43<br>(47.7) | 7<br>(7.7) | 1<br>(1.1) | 0 | 90 (1 no response) |
| Create interest in topic | TL | 43<br>(41.7) | 51<br>(49.5) | 4<br>(3.9) | 3<br>(2.9) | 2<br>(1.9) | 103 |
|  | FC | 49<br>(53.8) | 38<br>(41.8) | 4<br>(4.4) | 0 | 0 | 91 |
| Increase motivation to learn | TL | 37<br>(35.9) | 52<br>(50.5) | 10<br>(9.7) | 3<br>(2.9) | 1<br>(1) | 103 |
|  | FC | 42<br>(46.2) | 38<br>(41.8) | 11<br>(12.1) | 0 | 0 | 91 |
| Better organisation of topic | TL | 38<br>(37.2) | 47<br>(46.1) | 14<br>(13.7) | 3<br>(2.9) | 0 | 102 (1 no response) |
|  | FC | 42<br>(46.2) | 44<br>(48.4) | 5<br>(5.5) | 0 | 0 | 91 |
| Generate peer interaction | TL | 38<br>(36.9) | 43<br>(41.7) | 17<br>(16.5) | 5<br>(4.9) | 0 | 103 |
|  | FC | 47<br>(51.6) | 38<br>(41.8) | 6<br>(6.6) | 0 | 0 | 91 |
| Promote team work | TL | 41<br>(39.8) | 38<br>(36.9) | 18<br>(17.5) | 6<br>(5.8) | 0 | 103 |
|  | FC | 47<br>(51.6) | 38<br>(41.8) | 6<br>(6.6) | 0 | 0 | 91 |
| Recommend to juniors | TL | 53<br>(51.5) | 34<br>(33) | 10<br>(9.7) | 2<br>(1.9) | 4<br>(3.9) | 103 |
|  | FC | 43<br>(47.7) | 42<br>(46.6) | 5<br>(5.5) | 0 | 0 | 90(1 no response) |
| Facilitate teacher student interaction | TL | 48<br>(46.6) | 49<br>(47.6) | 6<br>(5.8) | 0 | 0 | 103 |
|  | FC | 58<br>(63.7) | 30<br>(33) | 3<br>(3.3) | 0 | 0 | 91 |
| Greater understanding of topic | TL | 48<br>(47.5) | 49<br>(48.5) | 2<br>(1.9) | 2<br>(1.9) | 0 | 101(2 no response) |
|  | FC | 47<br>(51.6) | 43<br>(47.3) | 1<br>(1.12) | 0 | 0 | 91 |

There was an overall perception that FC can enhance communication skills and provides a better organisation of the topic to be studied, the difference was not statistically significant (p= 0.054).

For all the perceptions, FC had greater percentage of responses in category 1 i.e. *Strongly agree* when compared to TL; whereas the responses in category 2 i.e. *Agree* was more for TL. This denotes that more students favour FC versus TL though the difference was not much.

## Discussion

Medicine involves complex integration of various subjects and its clinical application aided by critical thinking. There is a need for self learning and life long learning to excel in this field. The FC method involves more interaction, facilitates critical thinking and is a learner centric approach which is an integral part of adult learning. As a result, FC has gathered a lot of backing from medical and related fields. The present study examined the acquisition of higher order cognitive skills assessed by prescription writing skills for clinical scenarios; as well as perception of students towards TL and FC on ten parameters. The marks obtained after evaluating the prescription writing exercise were not significantly different between the TL and FC groups, though the perception that academic score will increase with FC was there. A systematic review by Ramnanan CJ highlights the same aspect of FC i.e. eventhough the student response towards FC is largely positive, there is no convincing evidence that FC enhances learning.^9^ Another study which examined the FC approach to teaching evidence based medicine did not improve scores compared to its traditional counterpart.^10^ The reason for this result in our study may be due to the implementation of FC for just one session. Since the students had no previous experience of FC, they might have been a bit slow to adopt this new method. Another explanation why performance scores generally are not significantly different with FC compared to TL may due the testing methods followed which are not adequate in assessing the higher order cognitive skills.^11^

In this study, there was statistically significant difference in the perceptions of students in favour of FC with regards to ability to improve academic score, generate peer interaction, promote team work and facilitate teacher student interaction. Whereas such a significant difference was not seen in reference to ability to enhance communication skills, create interest in topic, increase motivation to learn, better organisation of topic or provide them with greater understanding of the topic and thereby not recommending FC over TL to their juniors. Flipped classroom is better than TL in acquiring Blooms higher order skills like analysis while for lower order skills like memorising and understanding FC is as good as TL.^12^ RA Blair et.al. looked at the knowledge gain as well change in prescription pattern for type 2 diabetes mellitus among internal medicine post-graduates and found that FC was preferred by the trainees. They also reported increase in knowledge as well as change in prescribing pattern following FC.^13^ Most studies on FC demonstrate greater student satisfaction which stems from the fact that students can learn at their own pace and at their own convenience. They also enjoy the small group discussions and problem-solving exercises during the face-to-face sessions.^14^ It has been suggested that FC model when implemented properly improves both intrinsic and extrinsic learner motivation.^15^ A quiz implemented at the beginning of the face-to-face session has been found to increase student motivation to go through the pre-class content and also helps in gauging student leaning. Tests can be conducted at various points of time like at the start as well as end of face-to-face session, after a few weeks to even months after the session. While gauging student learning, these tests also increase student scores by test-enhanced learning. Another advantage is that the tests help to evaluate the effectiveness of the teaching method.^16^ There is some objective evidence that FC enhances student interest and engagement as seen by the marked increase of attendance from 30% to 80% when lecture classes were flipped.^17^ The learner centric approach of FC has resulted in increased peer interaction and communication during the pre-class review of study materials as well as during the small group discussions and problem-solving sessions. The teacher acts as a facilitator during the face-to-face sessions providing support and a framework as the students progress towards their learning objectives. The FC method facilitates greater teaches student interaction and peer interaction when compared to TL.^18^

Students of medical and allied fields are generally appreciative of the FC method, though they have their apprehensions.^9^ There is a need for student to spent in the pre-class phase since there is the need to go through the topic related content which is time intensive. The preparatory material may not be appropriately aligned to the content covered in the classroom sessions. Some faculty may be overly didactic in the face-to-face sessions which can negate the benefits of FC. Unless the faculty facilitate discussion by all students in each small group, some students may dominate the sessions. These limitations of the FC can be overcome by proper planning and improving the content and method after collecting feedback from students. Philips J has in a short communication described an excellent framework for implementation of the FC model as well as given a checklist for the best practices.^19^

## Conclusion

Due to change in curriculum, there is less formal time for teaching pharmacology. There is also increased emphasis of self directed learning and small group discussions. The FC method in addition to engaging students in an active and collaborative learning process, is also valuable in introducing students to important topics in the subject outside of the formal curricular time. While there is a generally favourable mindset of students towards flipped classroom, there was no significant improvement in academic performance. Studies which involve the students for a greater length of time in the flipped classroom method in Pharmacology are needed.

## Data Availability

All data produced in the present study are available upon reasonable request to the authors. All results analyzed from the data are contained in the manuscript.

